# Noncommunicable Diseases, Sociodemographic Vulnerability, and the Risk of Mortality in Hospitalized Children and Adolescents with COVID-19 in Brazil: A Syndemic in Play

**DOI:** 10.1101/2021.02.11.21251591

**Authors:** Braian Lucas Aguiar Sousa, Alexandra Valeria Maria Brentani, Cecilia Claudia Costa Ribeiro, Marisa Dolhnikoff, Sandra Josefina Ferraz Ellero Grisi, Ana Paula Scoleze Ferrer, Alexandre Archanjo Ferraro

## Abstract

**Background:** Although many studies identify the presence of comorbidities and socioeconomic vulnerabilities as risk factors for worse COVID-19 outcomes, few have addressed this issue in children. We aimed to study how these factors have impacted COVID-19 mortality in Brazilian children and adolescents.

**Methods:** This is an observational study using publicly available data from the Brazilian Ministry of Health. We studied 5,857 patients younger than 20 years old, all of them hospitalized with laboratory-confirmed COVID-19. We used multilevel mixed-effects generalized linear models to study mortality, stratifying the analysis by age, region of the country, presence of noncommunicable diseases, ethnicity, and socioeconomic development.

**Findings:** Individually, most of the comorbidities included were risk factors. Having more than one comorbidity increased almost tenfold the risk of death (OR 9·67 95%CI 6·89-13·57). Compared to White children, Indigenous, *Pardo* (mixed), and East Asian had a significantly higher risk of mortality. We also found a regional effect (higher mortality in the North), and a socioeconomic effect (higher mortality among children from less socioeconomically developed municipalities).

**Interpretation:** Besides the impact of comorbidities, we identified ethnic, regional, and socioeconomic effects shaping the mortality of children hospitalized with COVID-19 in Brazil. Putting these findings together, we propose that there is a syndemic among COVID-19 and noncommunicable diseases, driven and fostered by large-scale sociodemographic inequalities. Facing COVID-19 in Brazil must also include addressing these structural issues. Our findings also identify risk groups among children that should be prioritized for public health measures, such as vaccination.

**Funding:** None.

## Introduction

The Coronavirus Disease 2019 (COVID-19) pandemic is the most significant health challenge of the century, with more than 80 million infected and 1·8 million deaths as of December 2020.^1^ After a first wave that elicited radical containment measures around the world, during the last months of 2020, we have watched the numbers rise again, prompting countries to reinstate lockdowns and reinforce containment policies, with special focus on vaccination. Unfortunately, Brazil has lagged. The federal government has been widely criticized for questioning the seriousness of the disease or denying its gravity altogether, delaying a timely response.^2,3^ As of December 2020, Brazil ranks third in number of cases and second in deaths.^1^ The country is currently struggling with its vaccination strategy, an effort hindered by partisan politics.

Children and adolescents are mostly spared by COVID-19, with few having severe symptoms and even fewer dying.^4^ However, the description of the multisystem inflammatory syndrome in children reinforced that, although rare, severe clinical presentation and death is possible in the pediatric population.^5^ Multiple studies have associated the presence of underlying comorbidities with severe clinical presentation and unfavorable outcomes in pediatric COVID-19 patients,^6-8^ however this association is less established than for adults. Additionally, it’s well recognized that ethnic minorities and those with less favorable socioeconomic status (SES) suffer a disproportionate impact of the COVID-19 pandemic,^9,10^ but this relationship hasn’t yet been studied for children specifically.

To make sense of its interaction with noncommunicable diseases (NCDs) and sociodemographic vulnerabilities, COVID-19 has been proposed to be a part of a syndemic rather than a pandemic.^11^ The syndemic theory was developed in medical anthropology as a way to explain how diseases can interact with each other, clustering in specific unfavorable socioeconomic settings. It is based on the synergism of two or more health conditions and the underlying socioeconomic inequality context. The combination of the conditions involved in a syndemic worsens their individual health outcomes, moving beyond the multimorbidity model to study how the socioeconomic and cultural factors promote these interactions.^12,13^

In this study, we analyze a large dataset of COVID-19 hospitalized children and adolescents to assess risk factors for mortality in this age group. We focused our attention on the impact of NCDs and sociodemographic variables such as country region, socioeconomic development, ethnicity, and age.

## Methods

### Study design and population

This is a cross-section observational study using publicly available data from the Brazilian Ministry of Health. We analyzed the SIVEP-Gripe database,^14^ which contains prospectively-collected data from all patients with severe acute respiratory syndrome (SARS) across the country. In Brazil, the notification of SARS is mandatory, and all the registered cases are included in the dataset. The reporting form is standardized and usually filled in the setting of hospitalization. The studied data comprises all reported COVID-19 hospitalizations in Brazil, up to December 7^th^, 2020. (appendix pg.1)

Brazil is divided into 26 states and the Federal District, and grouped in 5 macroregions: North, Northeast, Central-west, Southeast, and South. For analytical purposes, we chose to divide the country into two maximally contrasting regions: North (comprising the North and Northeast macroregions) and South (comprising the Central-west, Southeast, and South macroregions). This division was based on economic, health, and educational indexes, and is usual in sociodemographic and economic studies of the Brazilian population. Previous literature on COVID-19 in Brazil also divided the country in a similar way.^15^

The Brazilian Institute of Geography and Statistics (IBGE) divides the Brazilian population in five categories, based on self-reported skin color: *Branco* (White), *Amarelo* (East Asian), *Preto* (Black), *Indígeno* (Indigenous), and *Pardo*.^16^ *Pardo* refers to mixed and diverse ethnic background, as a result of the intense miscegenation that characterizes Brazilian people. In late 2020, 46·4% of Brazilians identified themselves as *Pardo*, 44% as White, 8·6% as Black, and 1% as East Asian or Indigenous.^17^

GeoSES is a Brazilian composite SES index that incorporates education, poverty, mobility, wealth, segregation, income, and deprivation of resources and services, generating a score for each municipality ranging from −1 (less developed) to 1 (most developed) with good association with the Human Development Index.^18^ We used the GeoSES of the patient’s municipality as a proxy of SES. It is important to note that in Brazil there is a considerable overlap between the regional, ethnic, and socioeconomic variables mentioned above. (appendix pg.3)

To address NCDs, we retrieved data from the SIVEP-*Gripe* on previous comorbidities. The dataset comprises the following conditions: cardiovascular disease, hematologic disease, hepatic disease, asthma, diabetes, neurological disease, pulmonary disease, immunodepression, kidney disease, obesity, Down Syndrome, and “other comorbidities”. Therefore, most of the diseases are included in groups of diagnosis, rather than specific conditions.

The rate of missing data or data reported as “unknown” varied among the variables: 1,385 (23·6%) patients had no ethnicity recorded and 166 (2·8%) did not have GeoSES data available. Sex was unknown for one patient. For comorbidities, we chose to assume missing data as absence of that comorbidity. (appendix pg.2)

### Statistical analysis

Categorical variables were described using their absolute and relative frequency and continuous variables using their mean and standard deviation. Distribution according to outcome was initially analyzed through chi-square test (categorical variables), t-test (continuous variables normally distributed) or Kruskal-Wallis rank test (continuous variables not normally distributed).

Multilevel mixed-effects generalized linear models were built to calculate the odds ratio and 95% confidence intervals between exposure and outcome. We assumed the municipality where the subject resided and the health unit where hospitalization occurred as random effects. Crude analyses were followed by adjusted models for confounders (sex, age, GeoSES, ethnicity, and country region), and by models accounting for interaction. Given the broad age range in pediatrics, the final models were presented separately for those 10 years old or younger and older than 10 years old.

## Results

As of December 7th, the SIVEP-*Gripe* database included 1,000,024 patients. We filtered it by age to only include patients less than 20 years old, finding a total of 79,498. Of that total, only 7,706 had a positive PCR for SARS-CoV-2, and 758 of them were not hospitalized. Out of the 6,948 remaining, the outcome was known for 5,857, the final population included in our analysis (Figure 1).

**Figure 1:**
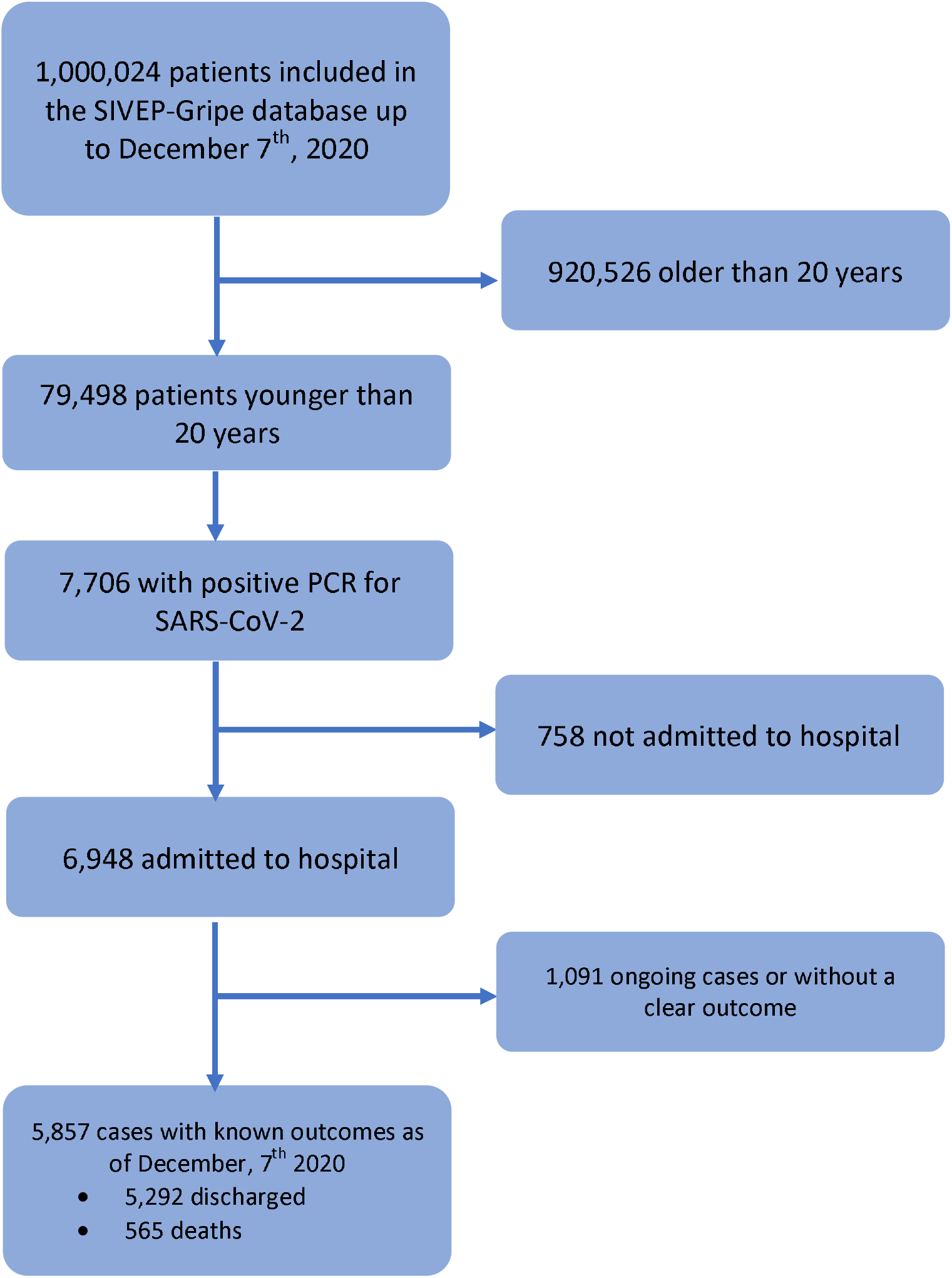
Flowchart of the patients included in this studied. PCR: polymerase chain reaction. SARS-CoV-2: Severe acure respiratory syndrome Coronavirus type 2.

Table 1 describes the sociodemographic and clinical features of the included patients. There was an even distribution among sexes, both in hospitalization (51·4% male vs 48·6% female) and in death rates. The mortality for age follows a U-shaped curve, with a higher death rate among neonates and adolescents.

**Table 1:** Sociodemographic description and preexisting noncommunicable diseases for survivors and non-survivors. Data are number (%) or mean (standard deviation). Missingness was found in both ethnic variables and GeoSES. One patient had no data on “sex”. NCD: noncommunicable disease.

| Characteristic | Categories | Survivors<br>(N=5,292) | Non-Survivors<br>(N=565) |
| --- | --- | --- | --- |
| Region | North (N=2,123) | 1,801 (84.8%) | 322 (15.2%) |
|  | South (N=3,734) | 3,491 (93.5%) | 243 (6.5%) |
| Age | 0-28 days (N=362) | 306 (84.5%) | 56 (15.5%) |
|  | 29d-2y (N=2,122) | 1,939 (91.4%) | 183 (8.6%) |
|  | 2y-10y (N=1,433) | 1,340 (93.5%) | 93 (6.5%) |
|  | 10y-15y (N=759) | 667 (87.9%) | 92 (12.2%) |
|  | 15y-20y (N=1,181) | 1,040 (88.1%) | 141 (11.9%) |
| Mean Age by Region<br>(Years) | North (N=2,123) | 6.6 (6.7) | 7.0 (7.2) |
|  | South (N=3,734) | 7.1 (6.9) | 9.0 (7.4) |
| Sex | Male (N=3,011) | 2,725 (90.5%) | 286 (9.5%) |
|  | Female (N=2,845) | 2,566 (90.2%) | 279 (9.8%) |
| Ethnicity | White (N=1833) | 1,704 (93%) | 129 (7%) |
|  | *Pardo* (N=2,363) | 2,070 (87.6%) | 293 (12.4%) |
|  | Black (N=199) | 182 (91.5%) | 17 (8.5%) |
|  | East Asian (N=36) | 30 (83.3%) | 6 (16.7%) |
|  | Indigenous (N=41) | 28 (68.3%) | 13 (31.7%) |
|  | Missing/Unknown Ethnicity<br>(N=1,385) | 1,278 (92.3%) | 107 (7.7%) |
| NCDs | With any NCD (N=2,318) | 1,952 (84.2%) | 366 (15.8%) |
|  | Without NCD (N=3,539) | 3,340 (94.4%) | 199 (5.6%) |
|  | Cardiovascular Disease (N=232) | 171 (73.7%) | 61 (26.3%) |
|  | Asthma (N=455) | 436 (95.8%) | 19 (4.3%) |
|  | Diabetes (N=150) | 118 (76.7%) | 32 (21.3%) |
|  | Pulmonary Disease (N=131) | 111 (84.7%) | 20 (15.3%) |
|  | Obesity (N=88) | 72 (81.8%) | 16 (18.2%) |
|  | Immunodepression (N=317) | 241 (76%) | 76 (24%) |
|  | Nerological Disease (N=299) | 243 (81.3%) | 56 (18.7%) |
|  | Renal Disease (N=96) | 68 (70.8%) | 28 (29.2%) |
|  | Liver Disease (N=32) | 25 (78.1%) | 7 (21.9%) |
|  | Hematologic Disease (N=149) | 123 (82.5%) | 26 (17.5%) |
| GeoSES |  | -0.058 (0.31) | -0.23 (0.33) |

The North region accounted for 36% of the included patients, the same proportion of the population living in the region. However, despite accounting for only one third of the hospitalizations, this region concentrated 57% of the deaths. We found an overall mortality of 9·6% in the country, but it varied among regions, with a higher rate in the North. We also found that although the distribution of cases among ethnicities roughly mirrors the population distribution, some ethnicities have a higher mortality than others, notably Indigenous and *Pardo*. Additionally, children who died lived in municipalities less socioeconomically developed than those who survived (GeoSES −0·23 *vs* −0·058, p<0·001).

In order to provide a clear picture of the disease distribution, we analyzed the number of hospitalizations and mortality state by state (Figure 2). São Paulo, the state with the largest population, leads with 1,880 reported hospitalizations. However, proportionally to the total state population, Sao Paulo was only the 4^th^ in the country, after Sergipe, Pernambuco, and the Federal District. Roraima, the least populated state in Brazil, had the highest mortality rate (70·6%), a figure that needs to be interpreted with care due to the low number of cases reported in the state. The states with the highest mortality rates were mostly concentrated in the North, which corroborates our approach of dividing Brazil into two regions.

**Figure 2:**
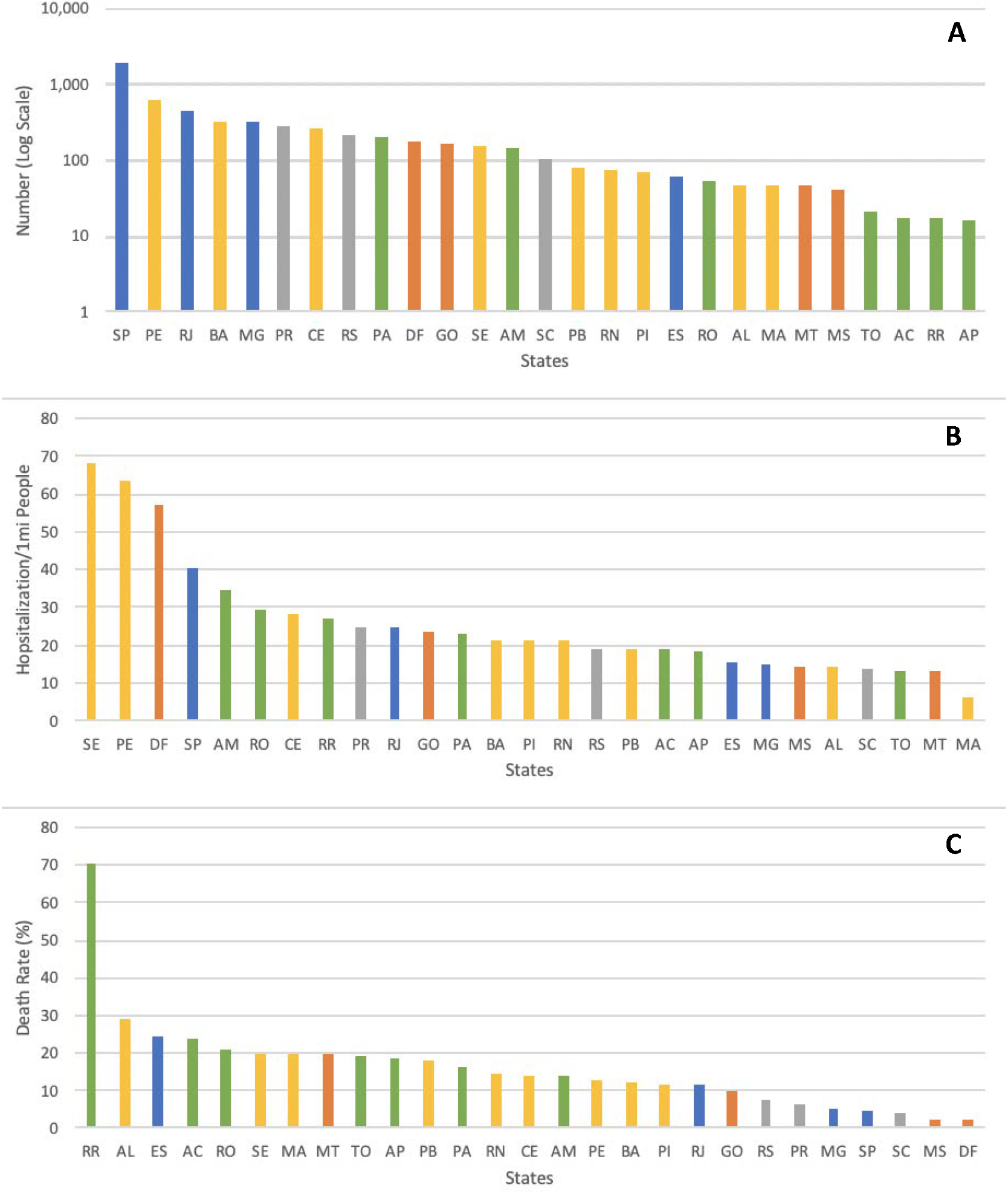
Distribution of COVID-19 pediatric hospitalizations in absolute numbers (A), proportional to population (B) and death rate (C) by state. N=5,857. Bars are colored by state macroregion: blue for Southeast; yellow for Northeast; grey for South; green for North; orange for Center west. AC=Acre. AL=Alagoas. AM=Amazonas. AP=Amapá. BA=Bahia. CE=Ceará. DF=Distrito Federal. ES=Espírito Santo. GO=Goiás. MA=Maranhão. MG=Minas Gerais. MS=Mato Grosso do Sul. MT=Mato Grosso. PA=Pará. PB=Paraíba. PE=Pernambuco. PI=Piauí. PR=Paraná. RJ=Rio de Janeiro. RN=Rio Grande do Norte. RO=Rondônia. RR=Roraima. RS=Rio Grande do Sul. SC=Santa Catarina. SE=Sergipe. SP=São Paulo. TO=Tocantins.

Analyzing NCDs prevalence, 39·6% of the children had at least one comorbidity, in the overall studied population and for both regions equally. The most frequently reported NCDs among hospitalized children with COVID-19 were asthma (7·8%) followed by immunodepression (5·4%), and neurologic conditions (5·1%). Multiple NCDs were reported by 10·4%. The overall mortality rate for children with NCDs was 15·8%, against 5·6% in healthy children. With the exception of asthma, all comorbidities increased the rate of death, especially kidney disease (29.2%) and cardiovascular disease (26.3%).

Assessing the odds ratio (OR) of mortality for all comorbidities and for cumulative comorbidities, we found that almost all categories significantly increased the risk of death. The exceptions were asthma, which appeared to be a protective factor (OR 0.42) in this population, and hepatic diseases. Individually, cardiovascular disease was the category that presented the highest effect on mortality (OR 4.98), followed by kidney disease, and immunosuppression. Having two or more comorbidities increased almost tenfold the odds of death (Figure 3). It is important to note that we excluded asthma when analyzing multiple comorbidities, since it’s protective effect would reduce the magnitude of the association.

**Figure 3:**
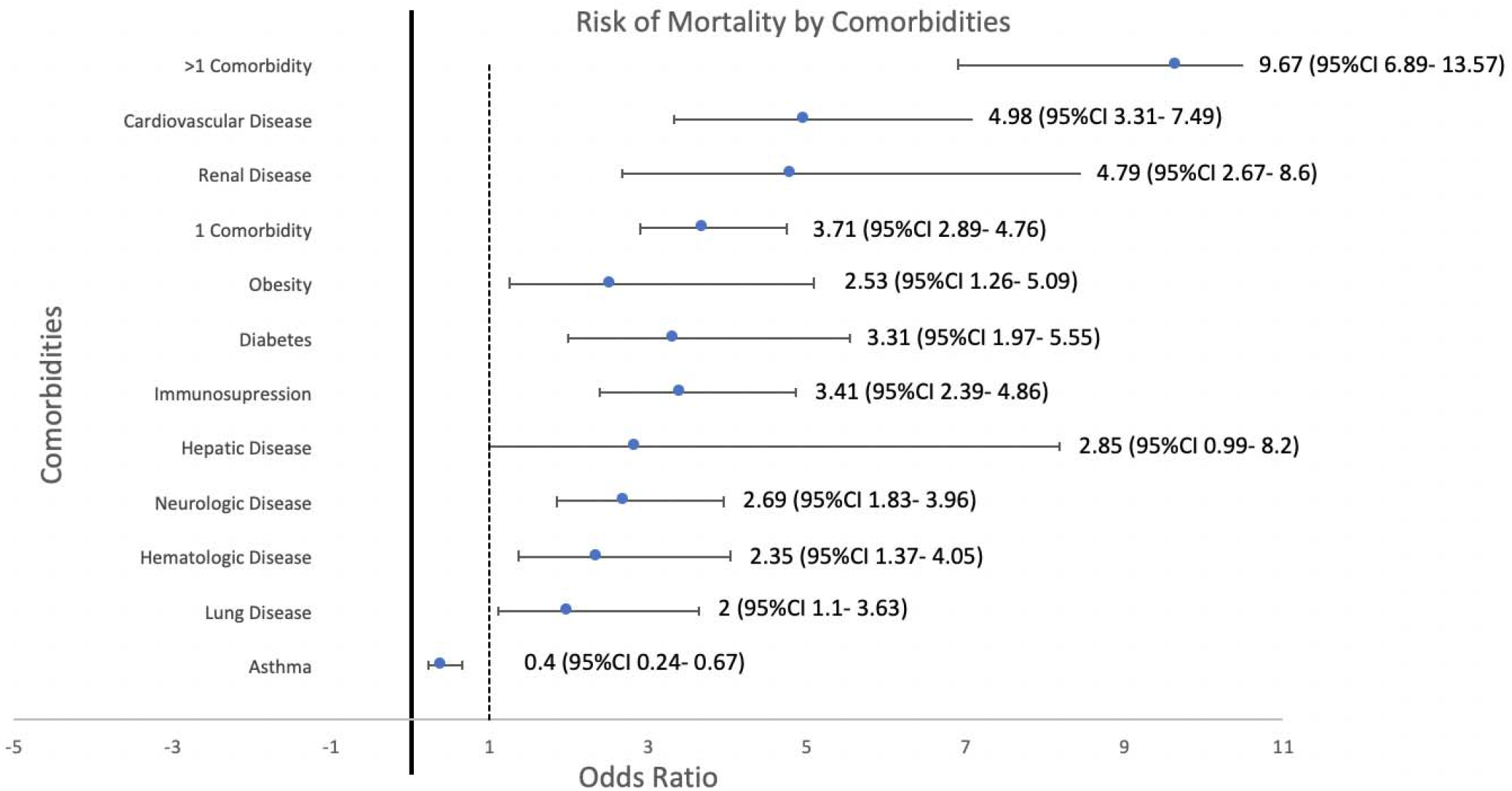
Risk of mortality for clinical features in multilevel mixed-effects generalized linear models, assuming municipalities and hospitals as random effects. Error bars represent 95% confidence intervals, N=5,857. For each individual condition, the reference group was patients without such condition. The groups “1 comorbidity” and “>1 comorbidities” includes all the aforementioned, Down syndrome and conditions disclosed as “other comorbidities”. Both categories exclude asthma, as asthma seems to has a protective effect. The reference group for them was patients without comorbidities.

Our next step was to study how regionality, socioeconomic, and ethnic variables impact mortality. Figure 4 illustrates the association between each of those factors and COVID-19 mortality in hospitalized children. We found that *Pardo* (OR 1·93), East Asian (OR 2·98) and Indigenous ethnicities (OR 5·83) have significantly increased mortality figures when compared to White. Analyzing regions, children in the North region had more than three times the risk of death when compared to those in the South. SES, estimated by the GeoSES index, also showed a significant correlation with mortality: children from cities in the middle third GeoSES group had a 50% lower risk of mortality when compared to those in the lowest third. As for those in more developed cities, the risk reduction was almost 75%.

**Figure 4:**
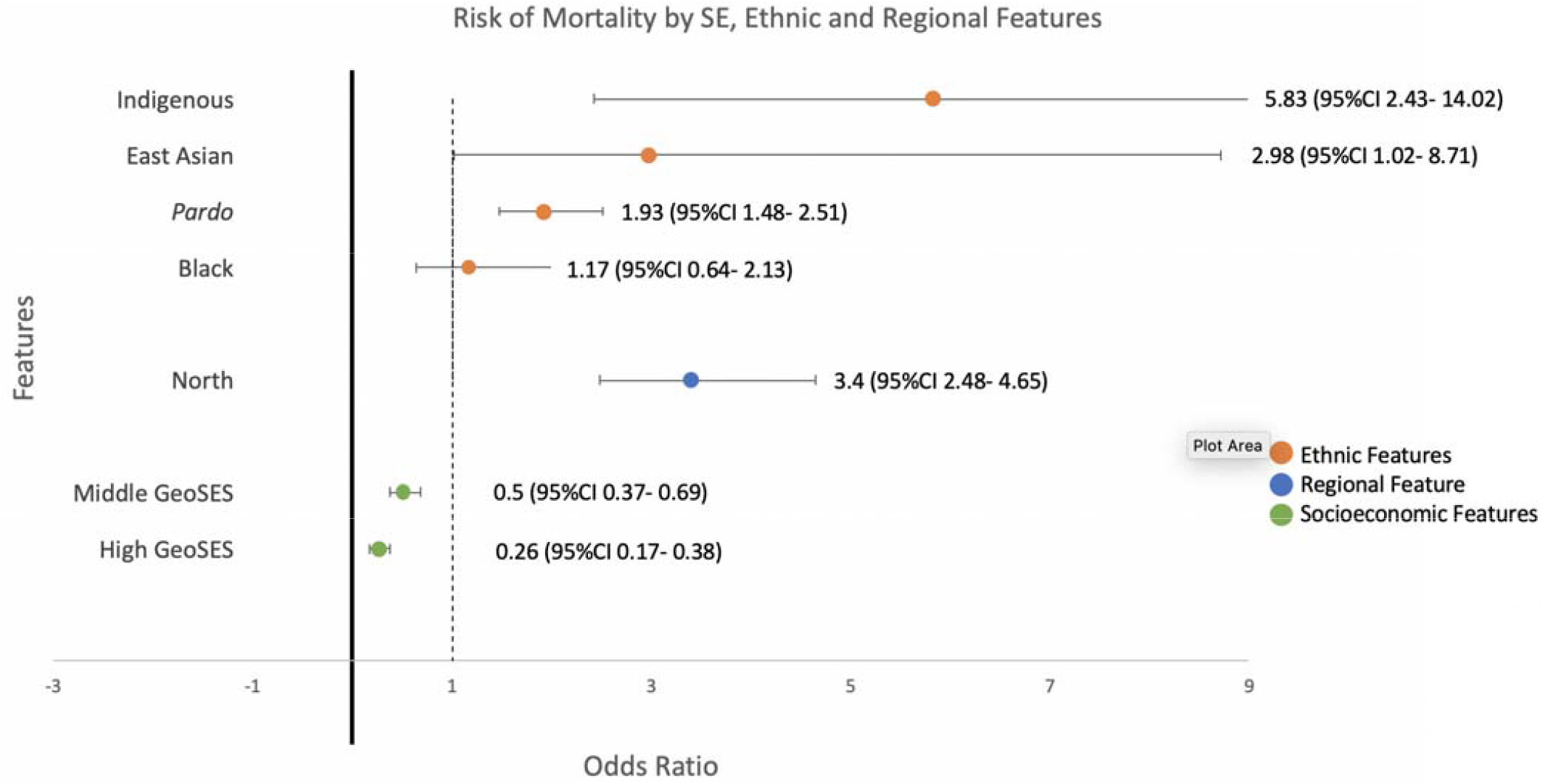
Risk of mortality for sociodemographic features in multilevel mixed-effects generalized linear models, assuming municipalities and hospitals as random effects. Error bars represent 95% confidence intervals. N=4,472 for ethnicity (1,385 missing), 5,857 for region, and 5,691 for socioeconomic development (166 missing). “*Pardo*” means a wide range of mixed or diverse ethnic backgrounds. “North” refers to both North and Northeast Brazilian macroregions. Socioeconomic development categorized by GeoSES terciles. Reference groups were “white” for ethnicity, “south” for region and Low GeoSES tercile for socioeconomic development.

To study the impact of all the aforementioned factors together on mortality, we developed five models (Table 2). Model 1 accounts for the impact of having comorbidities (at least one – asthma excluded) on mortality, adjusted for sex, and found an increase in the risk of death by almost five times. Adjusting for sociodemographic variables (age group, ethnic, region, and socioeconomic development – Model 2), similar results were found. Considering the possibility of interaction between comorbidities and the sociodemographic factors (Model 3), the impact of comorbidities in mortality is lower, but all the factors keep significance. Expanding the analysis, we split the patients into two age groups: 10 years old or younger (Model 4), and older than 10 years (Model 5). While the impact of NCDs appear to be more relevant for adolescents, in children the sociodemographic factors are more important. Moreover, in adolescents sex seems to be a relevant feature, with females having a 30% reduction in the risk of death, although the 95% CI included the unity (p=0.055).

**Table 2:** Explanatory models for the relationship between sociodemographic factors and mortality in hospitalized children with COVID-19 in Brazil. Results of multilevel mixed-effects generalized linear models, with municipality and hospital as random effects. Model 1 considers exclusively the impact of comorbidities (at least one, asthma excluded) in mortality, adjusted for sex. Model 2 includes adjustment for sociodemographic factors (ethnicity, region, age and GeoSES). Model 3 expands Model 2, including the assessment of possible interactions between comorbidities and sociodemographic factors. Models 4 and 5 are equivalent to Model 3, filtering for age group (up to ten years old or older than ten years old, respectively). The reference category for “comorbidity” was patients without previous conditions. OR: odds ratio. 95%CI: 95% confidence interval.

| **Models** | **OR** | **95%CI** | **p** |
| --- | --- | --- | --- |
| **Model 1** |  |  |  |
| Comorbidity | 4.79 | 3.52-6.52 | <0.001 |
| **Model 2** |  |  |  |
| Comorbidity | 4.71 | 3.63-6.11 | <0.001 |
| **Model 3** |  |  |  |
| Comorbidity | 2.89 | 1.02-8.22 | 0.046 |
| White | 1.00 | - | - |
| Pardo | 2.13 | 1.29-3.54 | 0.003 |
| Black | 1.44 | 0.54-3.85 | 0.460 |
| Asian | 4.68 | 1.07-20.49 | 0.040 |
| Indigenous | 9.32 | 3.01-28.84 | <0.001 |
| South | 1.00 | - | - |
| North | 1.76 | 1.08-2.86 | 0.023 |
| Low GeoSES | 1.00 | - | - |
| Middle GeoSES | 0.51 | 0.31-0.81 | 0.005 |
| High GeoSES | 0.34 | 0.17-0.69 | 0.002 |
| Male | 1.00 | - | - |
| Female | 0.99 | 0.79-1.25 | 0.959 |
| **Model 4 (<=10yo)** |  |  |  |
| Comorbidity | 2.85 | 0.87-9.34 | 0.083 |
| White | 1.00 | - | - |
| Pardo | 2.17 | 1.14-4.11 | 0.018 |
| Black | 2.13 | 0.63-7.17 | 0.220 |
| Asian | 5.79 | 0.92-36.30 | 0.061 |
| Indigenous | 15.78 | 4.18-59.54 | <0.001 |
| South | 1.00 | - | - |
| North | 1.76 | 0.95-3.25 | 0.070 |
| Low GeoSES | 1.00 | - | - |
| Middle GeoSES | 0.58 | 0.30-1.12 | 0.105 |
| High GeoSES | 0.27 | 0.13-0.59 | 0.001 |
| Male | 1.00 | - | - |
| Female | 1.22 | 0.90-1.66 | 0.195 |
| **Model 5 (>10yo)** |  |  |  |
| Comorbidity | 5.33 | 1.55-18.27 | 0.008 |
| White | 1.00 | - | - |
| Pardo | 2.22 | 0.94-5.20 | 0.067 |
| Black | 0.88 | 0.16-4.78 | 0.880 |
| Asian | 3.04 | 0.24-39.15 | 0.393 |
| Indigenous | 2.18 | 0.13-35.38 | <0.584 |
| South | 1.00 | - | - |
| North | 1.70 | 0.74-3.89 | 0.206 |
| Low GeoSES | 1.00 | - | - |
| Middle GeoSES | 0.49 | 0.23-1.06 | 0.071 |
| High GeoSES | 0.35 | 0.12-0.97 | 0.043 |
| Male | 1.00 | - | - |
| Female | 0.68 | 0.46-1.00 | 0.055 |

Finally, we compared the rate of NCDs in the studied population to the rate among the deceased, for each sociodemographic category (Figure 5). It is clearly noticeable that in the North region, in lower socioeconomic settings, and among ethnic minorities, a higher proportion of healthy children died, even though the proportion of hospitalized children without comorbidities is fairly similar for all categories. This is also true for the younger than 10 years, as in adolescence more than 70% of the deaths were in children with at least one NCD. However, in the age categories we see a larger gap in NCDs prevalence, with a higher rate among adolescents.

**Figure 5:**
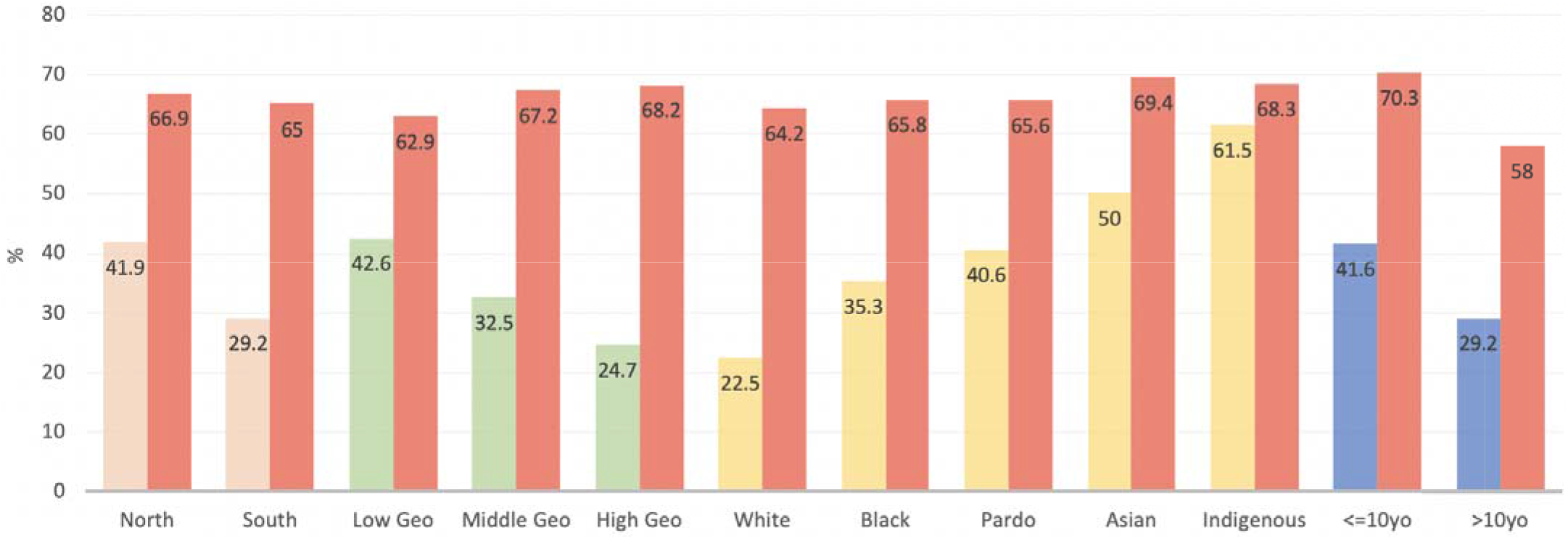
Frequency of children without NCDs for each sociodemographic category, both in the general hospitalized population and among the dead. Bars in red represent the general rate for each group, while the other bars represent the rate among the dead. “*Pardo*” means a wide range of mixed or diverse ethnic backgrounds. “North” refers to both North and Northeast Brazilian macroregions. Socioeconomic development categorized by GeoSES terciles.

## Discussion

This is, to our knowledge, the most comprehensive study on risk factors for mortality of hospitalized children with COVID-19, covering both clinical and sociodemographic characteristics in a large population. We described the higher risk of mortality associated with having NCDs as well as among ethnic groups such as Indigenous and *Pardo*. We also found regional and socioeconomic disparities associated with mortality in this specific population, painting a broad picture of how these sociodemographic elements interact with NCDs to shape mortality in COVID-19 hospitalized children.

We found significant differences in mortality rate when splitting age in groups to consider previously described ages of potential risk, like neonates and older adolescents.^19^ The higher mortality in neonates might be related to the developing immune system, associated with an immature respiratory system, resulting in them being more prone to severe complications of lung infections. Adolescents, in turn, bear a higher burden of NCDs. Accordingly, in our explanatory models, having at least one NCD increased the risk of death over five times for adolescents, while for those younger than 10 years, the comorbidity effect was not significant.

The proportion of hospitalized children among the regions was consistent with the national population, evidencing a uniform distribution of the disease. As we studied data up to December, our results mirror a late stage of the pandemic in Brazil, with a widespread distribution of COVID-19. The state reporting the highest number of hospitalized children with COVID-19 was Sao Paulo. However, proportionally to population size, Sergipe, a small state in the Northeast macroregion, with 20 times less people than Sao Paulo, and significantly lower levels of socioeconomic development and health indicators, took the lead. These findings reinforce the idea that COVID-19 is spreading especially among more vulnerable populations, which can have catastrophic public health consequences. Two national serologic household surveys also found inequalities in the prevalence of the disease, with a higher prevalence in poorer areas and among minorities.^20^

We found a high prevalence of NCDs (39·6%) in hospitalized children with COVID-19, corroborating the findings of other studies.^6-8^ Almost all the conditions studied individually posed as risk factors for mortality, however, it was their association that stood out. Due to the medical advances of the last decades, especially in newborn care and NCD treatments, we have seen a significant rise in the prevalence of children with multiple chronic conditions and medical complexity.^21^ Our findings support the idea that this population is at a higher risk of mortality, and deserves special attention with respect to preventive measures, including vaccination.

Other than multiple comorbidities, indigenous ethnicity was the most important risk factor for mortality in the population studied. Brazil is a country marked by discrimination and governmental negligence against Indigenous populations, reflecting on shortcomings in multiple socioeconomic and health indexes. Mortality for Indigenous children and adolescents, for instance, is much higher than for non-Indigenous.^22^ This setting of structural disadvantage reflects on special vulnerability to diseases: in 2009, for example, the H1N1 influenza devastated Indigenous tribes, with a mortality 4·5 times higher than the general Brazilian population.^23^ The current Brazilian political scenario makes matters even worse: deforestation is on the rise, the National Indigenous Foundation was stripped of power, and illegal miners and loggers are infiltrating Indigenous territories without governmental challenge.^24^ Our data and previous literature^20,23^ raise the alarm to a significantly higher risk of COVID-19 spreading and mortality among Indigenous, prompting quick and decisive governmental intervention.

Besides Indigenous, we identified *Pardo* and East Asian ethnicities as risk factors for mortality. In the early stages of the pandemic in Brazil, Marra and colleagues also found *Pardo* ethnicity as a risk factor in adults, attributing it to a greater susceptibility of contracting COVID-19, reliance on public health care, and reduced access to intensive care.^15^ It is reasonable to assume that *Pardo* children are subject to the same difficulties as adults, although the assessment of pediatric intensive care unit availability is compromised by the lack of official data on the subject. Additionally, *Pardo* and Black children also have higher general mortality rates when compared to White,^22^ mirroring generations of structural socioeconomic and health disadvantages. Surprisingly, in our study, black ethnicity was not associated with a higher mortality. The higher risk for East Asians is consistent with the findings of a recent meta-analysis including more than 18 million patients of all ages.^25^

The GeoSES index incorporates multiple social and economic dimensions, including variables often neglected like mobility and segregation.^18^ It is, therefore, a more comprehensive proxy than the ones classically used, like the HDI. The analysis by GeoSES terciles clearly demonstrates the abyss separating different levels of privilege in Brazilian society. It is appalling to see children dying almost 4 times more in cities less socioeconomically developed. In the battle against COVID-19, these children are clearly being left behind.

We also found a regional effect, with a higher mortality in Northern Brazil. Even in models incorporating all the sociodemographic factors together, the correlation stands, showing that there is a regional outcome discrepancy that is not explained by ethnicity or socioeconomic development alone. Marra and colleagues also found this regional effect, attributing it to a difference in health care availability and variation in number of comorbidities.^15^ Since the North region is less developed than the South, with worse health indexes, it is reasonable to assume that health care is also less available for children. However, we could not find a clear difference in the prevalence of comorbidities among regions. This finding must be interpreted cautiously, as a lower availability of health care renders underdiagnosis more likely. We also expect that children in Northern Brazil have a worse control of their chronic conditions, contributing to poor outcomes.

While we found a fairly similar proportion of children without NCDs in both regions, among different ethnicities and in the GeoSES terciles, the rate among the deceased varies remarkably. There is a clear pattern of an excess of deaths in healthy children from less favorable socioeconomic settings and among minorities. While this might be related to underdiagnosis, it also raises an alarm for a possible failure of the Brazilian health system in protecting these populations.

Approaching the COVID-19 as a syndemic rather than a pandemic helps understanding some of the complex relations the disease has established with NCDs and sociodemographic features. We propose that there is a syndemic between COVID-19 and NCDs in Brazilian children, driven by large-scale socioeconomic forces that promote the concentration and interaction of these conditions. For a syndemic to occur, there must be two epidemics interacting. Our findings clearly show that the presence of NCDs worsens the outcomes of children with COVID-19, especially when multimorbidity is present.

We have evidence that the opposite is also true: COVID-19 can have an impact on people with NCDs, either increasing the risk of developing them or worsening their care. COVID-19, for instance, can impair glucose metabolism and complicate preexisting diabetes or even have a diabetogenic effect.^26^ COVID-19 preventive strategies are based primarily on reducing social contact, which might lead to increased exposure to NCDs risk factors, like tobacco use and sedentarism. Furthermore, lockdowns have disrupted health care access for patients living with chronic conditions, making disease management harder.^27^

Our data reinforces the previously described idea that COVID-19 is spreading mainly through vulnerable populations, with a higher prevalence in states in the Northern region. Many factors might explain this higher transmissibility, like crowded living conditions and poor access to public health measures and healthcare.^20,25^ Therefore, it is reasonable to conclude that the pandemic is clustering in vulnerable populations, driven by the major socioeconomic forces that are determinant to health. Literature shows that NCDs also cluster in these same populations, that are more exposed to risk factors like unhealthy diets, physical inactivity, and tobacco use. This is also true for children, with the exposure to these factors starting in the womb and continuing through life. The social determinants of health have a greater impact over children, since they are not able to advocate for themselves, and are socially and economically dependent on their caregivers.^28^

Approaching COVID-19 as part of a syndemic invites a broader vision that goes beyond biomedical solutions and encompasses the socioeconomic environment that promotes the disease cluster and interaction with NCDs(11). Adequate treatment, preventive measures, and vaccination are not enough: governmental intervention is necessary to address the challenge of changing disparities structurally rooted in Brazilian society. Fortunately, Brazil has precedent on this topic: the *Bolsa Familia*, a nationwide conditional cash transfer program that covers over 15 million families, has been shown to significantly reduce childhood mortality.^29^ This example illustrates how large governmental interventions tackling socioeconomic disparities can have a positive impact on children’s health.

Our study had several limitations. Our analysis relied on secondary data, with case ascertainment bias being a possibility. There was also a high rate of missingness for ethnicity, which could imbalance the results if the missingness was differential for some groups, but no evidence was found in literature to support this hypothesis. Ethnicity was defined on the basis of self-declared skin color or appearance, rather than ancestry, and there’s a significant overlap between the *Pardo* and Black categories. Underreporting is also an issue, especially in less advantageous socioeconomic contexts, which might have underestimated the effect size in our models. As for the SES analysis, using municipality development as a proxy for socioeconomic status can hide major discrepancies within each city, especially in large metropolises.

We were not able to fully address healthcare availability by ethnicity, socioeconomic status, and region, since our analysis was restricted to children only. Noticeably, the GeoSES index does not include a health component in its dimensions.^18^ Therefore, the different levels of socioeconomic status derived from the index do not cover health access or morbidity and mortality risks. We didn’t have data on out-of-hospital mortality, which might be substantial especially in lower socioeconomic settings, possibly resulting in an underestimation of the pandemic effect in these settings.

In conclusion, we have described how the presence of NCDs and sociodemographic vulnerabilities can impact the mortality of hospitalized children and adolescents with COVID-19 in Brazil. We have found a higher risk of death associated with most of the NCDs included, especially when more than one was present. Indigenous, *Pardo* and East Asian ethnicities, as well as the Northern region and lower socioeconomic development were also risk factors for mortality. Putting these findings together, we proposed a syndemic approach for COVID-19 and NCDs in Brazilian children. Our findings are relevant for public health policy makers, as the country is still planning its vaccination strategy and trying to find the best way to navigate the health challenges imposed by the COVID-19 pandemic.

## Data Availability

The study used publicly available data from the Brazilian Ministry of Health.

https://opendatasus.saude.gov.br/dataset/bd-srag-2020

