## Appendix for "Noncommunicable Diseases, Sociodemographic Vulnerability, and the Risk of Mortality in Hospitalized Children and Adolescents with COVID-19 in Brazil: A Syndemic in Play"

**THE SIVEP-GRIPE DATABANK**

Since the Influenza A (H1N1) pandemic in 2009, the Brazilian Ministry of Health performs active surveillance for Severe Acute Respiratory Syndrome (SARS) cases. SARS case notification is mandatory in Brazil, and the data is stored in the SIVEP-Gripe database, which comprises 155 features per entry, including age, ethnicity, municipality of residency, attending hospital, symptoms, comorbidities, investigation of etiology, treatment, outcomes, among others. The reporting form is standardized and usually filled in the setting of hospitalization, although not every patient included in the databank needs hospitalization. In the beginning of the COVID-19 pandemic, the disease was incorporated in the Influenza surveillance network, and all reported cases were included in SIVEP-Gripe. Data is continuously reviewed, as case investigation progresses. The defining criteria for Influenza syndrome and SARS have changed through the years, reflecting the current level of scientific knowledge and different epidemiological situations. The most recent definitions, in the light of the COVID-19 pandemic, are as follows^1^:

Influenza syndrome is defined as an acute respiratory illness with at least two of the following: fever, chills, sore throat, headache, cough, coryza, olfactory disturbances or gustatory disturbances. In children, nasal obstruction might also be included.

SARS is defined as patients with Influenza syndrome and:

- Dyspnea/respiratory discomfort OR
- Persistent thoracic pressure OR
- Peripheric oxygen saturation <95% OR
- Cyanosis
- In children, other signs might be considered, including nasal flaring, intercostal retractions, dehydration and inappetence.

**MISSING DATA**

The rate of missing data, or data reported as “unknown”, varied in the dataset for different variables. Table 1 shows reported numbers (and percentages) of patients with missing data for sex, ethnicity, age, and GeoSES. As already mentioned, the notification form is usually filled during hospitalization, with a bias towards the filling of positive information exclusively. Therefore, for noncommunicable diseases (NCDs), we assumed the absence of information as absence of that comorbidity.

Almost all the missing data for GeoSES (165 out of 166 cases) came from urban areas within the Federal District. This occurs because, contrary to the states, the Federal District does not contain municipalities, but administrative regions only. Therefore, the only place with GeoSES available is Brasilia, the first administrative region and the national capital. It is worth noting that, out of 174 hospitalized children with COVID-19 in the Federal District, only 9 were reported in Brasilia. The remaining cases were therefore excluded from the GeoSES analysis.

The high rate of missing/unknown for ethnicity might reflect the lack of importance given by healthcare personnel to data not concerning clinical features, as they seem less relevant for health outcomes. However, in a county plagued by structural ethnic inequalities, reporting ethnicity is vital to understand and fight these issues^2^. Patients that lacked data on ethnicity were excluded from the ethnicity analysis.

Data on “sex” was unknown for one patient.

| **Variable** | **Unknown** | **Missing** | **Total** | **% Missing/Unknown** |
| --- | --- | --- | --- | --- |
| Sex | 1 | 0 | 1 | 0·02 |
| GeoSES | - | 166 | 166 | 2·83% |
| Ethnicity | 1,106 | 279 | 1,385 | 23·6% |
| Age | 0 | 0 | 0 | 0 |

**Table 1: Frequency of missingness for the variables studied.**

**BRAZILIAN DEMOGRAPHICS**

Brazil is the fifth largest country in the world by area^4^ and the sixth by population, with 212 million people^5^. The country is divided in 26 states and the Federal District, and grouped in 5 macroregions: North (States of Acre, Amapá, Amazonas, Pará, Rondônia, Roraima, and Tocantins), Northeast (States of Alagoas, Bahia, Ceará, Maranhão, Paraíba, Pernambuco, Piauí, Rio Grande do Norte, and Sergipe), Central-west (States of Goiás, Mato Grosso, Mato Grosso do Sul and the Federal District), Southeast (States of Espírito Santo, Minas Gerais, São Paulo, and Rio de Janeiro), and South (States of Paraná, Rio Grande do Sul, and Santa Catarina). For analytical purposes, we chose to dichotomize the country in two maximally contrasting regions with respect to economic, health and educational indexes: North (macroregions North and Northeast) and South (macroregions Central-west, Southeast and South).

There is a significant intersection between socioeconomic development, ethnicity, and region. The GeoSES for the patients in the North region is significantly lower than for those living in the South region (Figure 1), showing an expected overlap between region and socioeconomic development. There is also an intersection between region and ethnicity, with a higher frequency of Whites in South and *Pardos* in North. To illustrate the racial distribution in Brazil, Patadata developed an interactive Brazilian map of racial distribution, using data from the 2010 Census and representing each person by one dot on the map and the ethnicities by color^3^. The map and the code is available for free. In Figure 2, we adapted the racial distribution map to include a line roughly dividing Brazil in North and South, allowing for a clear image of the ethnicity concentration in the country. Finally, ethnicity and socioeconomic status are also associated, as it is shown in figure 3.

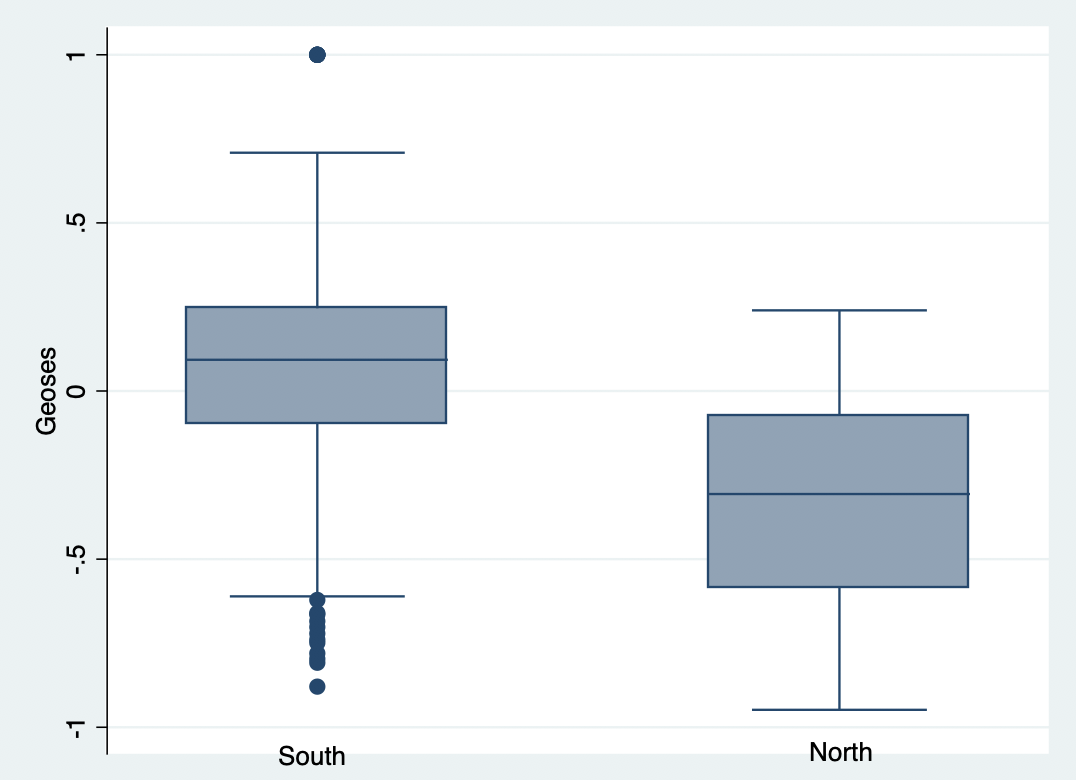

**Figure 1: Boxplot of the distribution of GeoSES by Region.** The mean Geoses is significantly different among regions (p<0·001).

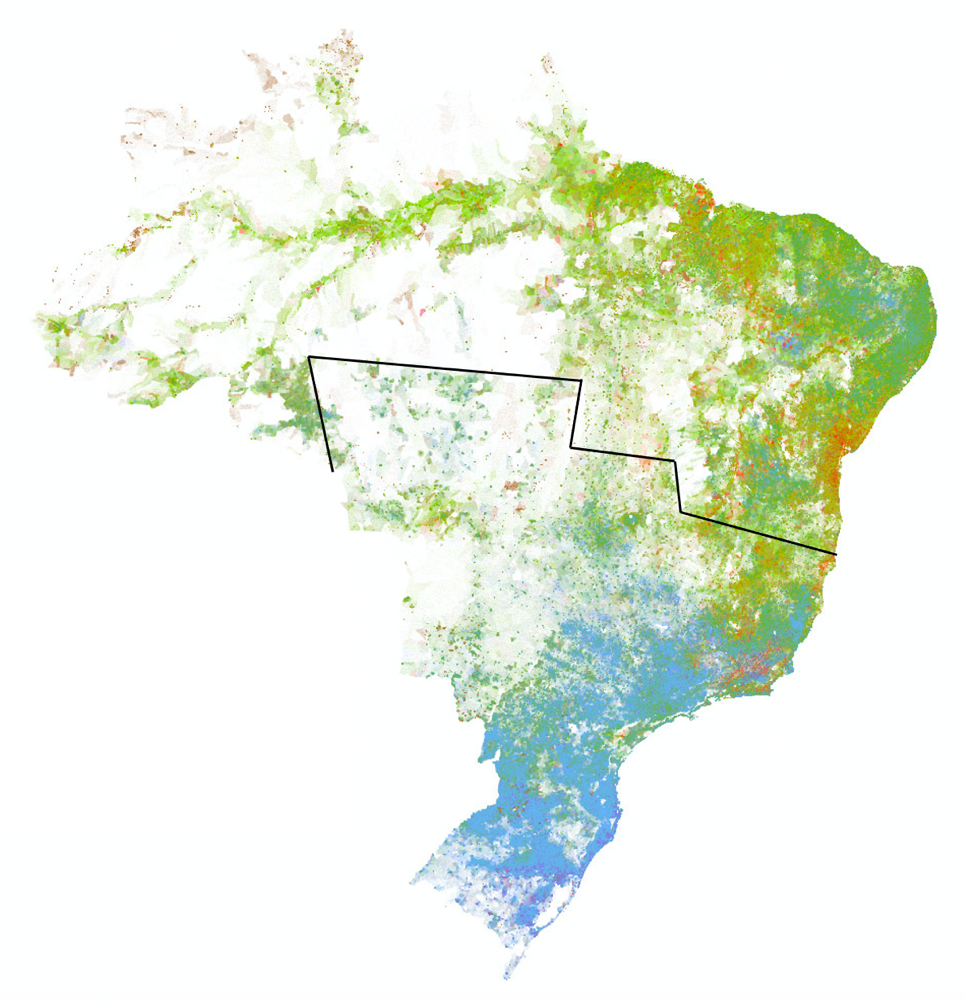

**Ethnicity**

Indigenous

East Asian

Black

*Pardo*

White

**Figure 2: Brazilian map of ethnic distribution according to the 2010 Census.** Adapted from the interactive map developed by Patadata^3^. The black line roughly divides Brazil in North and South. A concentration of *Pardos* is clearly visible in North, while in South, a higher frequency of Whites is observed.

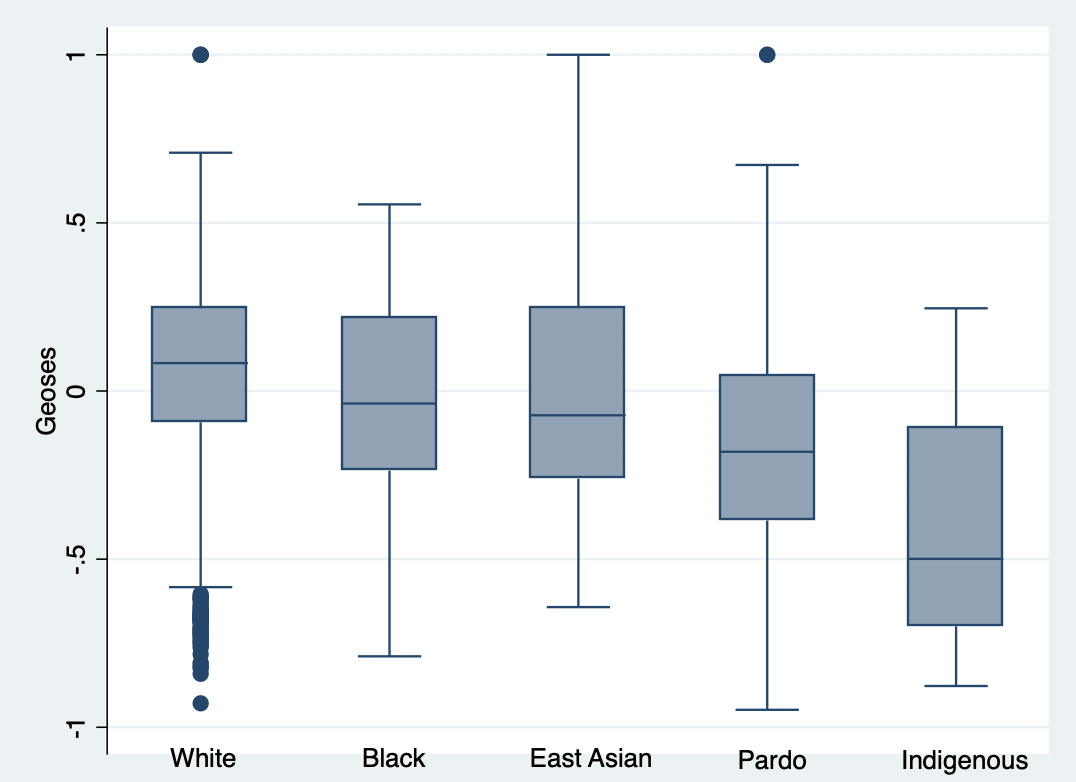

**Figure 3: Boxplot of the distribution of GeoSES by Ethnicity.**

**ADDITIONAL TABLE**

Table 2 describes the sociodemographic characteristics and preexisting NCDs for non-survivors by region. This table allows easily comparison of outcomes among regions for all the included variables.

|  | **Non-Survivors – NORTH** (N=322) | **Non-Survivors – SOUTH** (N=243) |
| --- | --- | --- |
| Age (years) | 7 (4/0·2-14) | 10 (10/0·6-17) |
| Male (N=1,106 S=1,905) | 172 (15·6%) | 114 (6%) |
| Female (N=1,017 S=1,828) | 150 (14·8%) | 129 (7·1%) |
| White (N=211 S=1,622) | 30 (14·2%) | 99 (6·1%) |
| Pardo (N=1,346 S=1,1017) | 214 (15·9%) | 79 (7·8%) |
| Black (N= 46 S=153) | 5 (10·9%) | 12 (7·8%) |
| East Asian (N=14 S=22) | 3 (21·4%) | 3 (13·6%) |
| Indigenous (N=26 S=15) | 9 (34·6%) | 4 (26·7%) |
| Missing/Unknown Ethnicity (N=480 S=905) | 61 (12·7%) | 46 (6%) |
| With any NCD (N=841 S=1,477) | 191 (22·7%) | 175 (11·9%) |
| Without NCD (N=1,282 S=2,257) | 131 (10·2%) | 68 (3%) |
| Cardiovascular Disease (N=83 S=149) | 27 (32·5%) | 34 (22·8%) |
| Asthma (N=124 S=331) | 8 (6·4%) | 11 (3·3%) |
| Diabetes (N=43 S=107) | 16 (37·2%) | 16 (15%) |
| Pulmonary Disease (N=30 S=101) | 7 (23·3%) | 13 (12·9%) |
| Obesity (N=10 S=78) | 2 (20%) | 14 (18%) |
| Immunodepression (N=143 S=174) | 36 (25·2%) | 40 (23%) |
| Nerological Disease (N=109 S=190) | 26 (23·9%) | 30 (15·8%) |
| Renal Disease (N=38 S=58) | 15 (39·5%) | 13 (22·4%) |
| Liver Disease (N=8 S=24) | 3 (37·5%) | 4 (16·7%) |
| Hematologic Disease (N=47 N=102) | 9 (17%) | 18 (17·7%) |
| GeoSES (N=1,866 S=3,027) | -0·39 (-0·45/-0·66 to -0·14) | -0·01 (-0·03/-0·2 to 0·21) |

**Table 2: Sociodemographic description and preexisting NCDs for non-survivors by region.** Data are number (%) or mean (median/interquartile range). Percentage of the total number of hospitalizations for each region. Missingness was found in both ethnic variables and GeoSES. One patient had no data on “sex”.
